# Alterations in CD39/CD73 Axis of T cells associated with COVID-19 severity

**DOI:** 10.1101/2021.09.18.21263782

**Authors:** Gilson P Dorneles, Paula C Teixeira, Igor M da Silva, Lucas L Schipper, Paulo C Santana Filho, Luiz Carlos Rodrigues Júnior, Cristina Bonorino, Alessandra Peres, Simone Gonçalves Fonseca, Marta Chagas Monteiro, Carina Rodrigues Boeck, Sarah Eller, Tiago F Oliveira, Eliana M Wendland, Pedro R T Romão

## Abstract

Purinergic signaling modulates immune function and is involved in the immunopathogenesis of several viral infections. This study aimed to investigate alterations in purinergic pathways in COVID-19 patients. Lower plasma ATP and adenosine levels were identified in mild and severe COVID-19 patients associated with proinflammatory cytokine profiles compared to healthy controls. Mild COVID-19 patients presented lower frequencies of CD4+CD25+CD39+ (activated/memory Treg) and CD4+CD25+CD39+CD73+ T cells, and increased frequencies of high differentiated (CD27-CD28-) CD8+T cells compared to health controls. Severe COVID-19 patients also showed higher frequencies of CD4+CD39+, CD4+CD25-CD39+ (memory T effector cell), high differentiated CD8+ T cells (CD27-CD28-) and diminished frequencies of CD4+CD73+, CD4+CD25+CD39+ mTreg, CD4+CD25+CD39+CD73+, CD8+CD73+ and low-differentiated CD8+ T cells (CD27+CD28+) in the blood in relation to mild COVID-19 patients and controls. Moreover, severe COVID-19 patients presented higher expression of PD-1 on low-differentiated CD8+ T cells. Both severe and mild COVID-19 patients presented higher frequencies of CD4+Annexin-V+ and CD8+Annexin-V+ T cells, showing increased T cell apoptosis. Plasma samples collected from severe COVID-19 patients were able to decrease the expression of CD73 on CD4+ and CD8+ T cells of a healthy donor. Interestingly, the in vitro incubation of PBMC from severe COVID-19 patients with adenosine reduced the NF-kB activation in T cells and monocytes. Together, these data add new knowledge regarding the immunopathology of COVID-19 through purinergic regulation, especially concerning adenosine deficiency.

**Brief Commentary:** *Background:* Host factors modulates the type and the strength of the immune response during the viral infection, as well as the disease outcomes. However, to date, the role of purinergic signaling in SARS-CoV-2 infection remains unclear. We sought to evaluate alterations in extracellular adenine nucleotides and CD39/CD73 axis in T cells and their relationship with acute COVID-19 immunopathogenesis.

*Translational Significance:* COVID-19 patients present lower extracellular ATP and adenosine levels associated with altered CD39 and CD73 expression in CD4+ and CD8+ T cells. Purinergic signaling correlated with alterations in the differentiation status of CD8+ T cells, lymphocyte mitochondrial membrane polarization and T cell apoptosis. Our demonstration of the lower NF-κB activation in T cells and monocytes after *in vitro* adenosine treatment may indicate the regulatory effect of adenosine in the inflammation and cytokine storm of COVID-19. This study adds new knowledge regarding the immunopathology of COVID-19 through purinergic regulation.

## 1. Introduction

The disease caused by SARS-CoV-2 (COVID-19) is an hyperinflammatory disease, that can be asymptomatic or manifested as a broad spectrum of disease ranging from few symptoms (mild COVID-19 cases) to severe pneumonia that may evolve to SARS and death (severe COVID-19 cases) (1). Some typical clinical symptoms of patients with COVID-19 are fatigue, fever, dry cough, dyspnea, and shortness of breath. Although a higher number of SARS-CoV-2 infections generate asymptomatic or mild COVID-19, while a proportion of the remaining cases show severe and critical pneumonia requiring oxygen support and mechanical ventilation (2).

Host factors and the physiological environment determine the type and the strength of the immune response during the viral infection, as well as the disease outcomes. Thus, the biology of several immune cells, including lymphocytes, can be influenced by blood extracellular adenine nucleotides – adenosine triphosphate (ATP), adenosine diphosphate (ADP), and adenosine monophosphate (AMP) and nucleoside (adenosine) (3). The purinergic signaling regulates a number or immune cell functions, such as cell-to-cell interactions, cell death, cytokine and chemokine secretion, surface antigen shedding, and cell proliferation (4). During infections, once released by damaged cells, ATP acts as a damage-associated molecular pattern through P2 receptors (P2X and P2Y receptors) activating immune cells and inducing strong pro-inflammatory effects (5). Interestingly, severe COVID-19 patients had higher ATP content in the bronchoalveolar lavage supernatant associated with P2RX7-inflammasome activation in macrophage compared to non-COVID-19 patients (6). Thus, it is possible that alterations in adenine-based purine molecules may contribute to the hyper inflammation and immune dysfunction observed in COVID-19 pathophysiology.

The ectoenzyme CD39 (ecto-nucleoside triphosphate diphosphohydrolase 1, E-NTPDase1) converts extracellular ATP into AMP, and then CD73 (ecto-5’-nucleotidase, E-5’NTase) dephosphorylates AMP into adenosine (4,7). Adenosine signaling is mediated by G-protein-coupled adenosine receptors (AR) which acts as a mechanism for regulating intracellular cyclic AMP (cAMP) levels to induce immunosuppressive events (8). CD39 and CD73 are present in several immune cells, such as CD4+ T cells (including regulatory T cells and effector memory T cells), CD8+ T cells, CD19+ B cells, and Natural Killer cells, and participate in the regulation of the duration and magnitude of the immune response (4). The expression and activity of both CD39 and CD73 changes under the pathological context of acute and chronic viral infections (9– 11). In addition, CD39 expression in CD8+ T cell has also been described as a marker of exhaustion in virus infection (12).

To date, Ahmadi and colleagues (13) found lower proportions of CD8+CD73-T cells in the peripheral blood of COVID-19 patients and demonstrated that this cells possess a significantly higher cytotoxic effector phenotype (13). In addition, CD4+ and CD8+ T cells from severe COVID-19 patients present a dysregulated status of activation, characterized by higher expression of human leukocyte antigen-DR (HLA-DR) and CD38 activation marker, associated with phenotypical and functional T cell exhaustion (14,15). To better understand the role of CD39/CD73 pathway in the immunopathology of SARS-CoV-2 infection, this study evaluated the systemic levels of ATP, ADP, AMP and adenosine and the frequencies of CD4+ and CD8+ T cells expressing CD39 and CD73 ectonucleotidases, the proportions of CD8+CD27^-/+^CD28^-/+^ expressing PD-1, as well as the rates of lymphocyte apoptosis in the peripheral blood from mild and severe COVID-19 patients.

## 2. Methods

### 2.1 Study patients and Ethics

We prospectively evaluated a convenience sample of mild and severe COVID-19 positive patients admitted at the Hospital Moinhos de Vento between July/2020 and November/2020 and volunteering uninfected healthy individuals. Infection with SARS-CoV-2 was confirmed by reverse transcription polymerase chain reaction (RT-PCR) with nasopharyngeal and oropharyngeal swab samples. This study was approved by the Moinhos de Vento Ethics Committee N 3977144 (Porto Alegre/Brazil) and informed consent was obtained from all participants. Blood samples (5 mL) were obtained from patients with mild or severe COVID-19 into K2EDTA tubes (Becton & Dickinson, USA) within 6 h from hospital admission. Flow cytometry experiments (described below) were conducted with fresh blood. The remaining blood was centrifuged (1500 *g*, 10 min), plasma was aliquoted and kept at −80 ºC until plasma analysis.

Disease severity was classified according to the World Health Organization classification after completing the follow-up questionnaire (16). Clinical and socio-demographic data were collected from the patient’s electronic medical records upon admission to the unit. Body mass index was calculated from weight and height data.

### 2.2 Analysis of adenine-based purine concentrations and lipopolysaccharide levels by liquid chromatography-tandem mass spectrometry (LC-MS/MS)

An aliquot of 150 μL of acetonitrile was added to the plasma samples (50 μL), and the mixture was shaken for 60 seconds. After centrifugation for 6 min at 9000 g, the supernatant was collected and a 25 μL aliquot was directly injected into the LC□ MS/MS system. The analytical system consisted of a Nexera UFLC system coupled to a LCMS-8040 triple quadrupole mass spectrometer (Shimadzu, Kyoto, Japan). The ESI-MS/MS parameters were set in positive and negative ion mode (polarity switch, 25 msec) as follows: capillary voltage, positive 4500 V and negative 3000 V; desolvation line temperature, 200 °C; heating block temperature, 500 °C; drying gas, 18 L/min; and nebulizing gas, 2 L/min. Analyses were carried out with multiple reaction monitoring (MRM) by using the following fragmentations: *m/z* 268.0□→□ *m/z* 136.0 for detection of adenosine ([M+H]^+^); *m/z* 346.0□→□ *m/z* 210.1 for detection of AMP ([M-H]^-^); *m/z* 426.0□→□ *m/z* 327.4 for detection of ADP ([M-H]^-^); and *m/z* 506.0□→□ *m/z* 207.5 for detection of ATP ([M-H]^-^). The chromatographic separation was conducted with a Shim-pack GISS column (2.1 × 100 mm, 1.9 μm particle size) (Shimadzu, Kyoto, Japan) eluted with flow rate of 0.3 mL/min. The gradient mobile phase system consisted of water (solvent A) and acetonitrile (solvent B) both fortified with 0.2% acetic acid and 0.1% tributylamine as follow: 0 – 4.5 min, 10 – 90% of B; 4.5 – 5.5 min, 90% of B; 5.5 – 5.6 min, 90 – 10% of B; 5.6 – 10 min, 10% B. The column oven was kept at 30 °C. The data were processed using LabSolutions software (Shimadzu, Kyoto, Japan). In addition, LPS concentration was determined by quantitation of 3-hydroxytetradecanoic acid as described by Teixeira and coworkers (17).

### 2.3 Systemic Cytokine levels

The plasma concentrations of IL□17A (from PeproTech, USA), and IL□6, IL□10 and TGF-β (all from eBioscience, ThermoFisher, USA), were quantified by enzyme□linked immunosorbent assay (ELISA) in microplate reader (EzReader, EUA). The detection limits of each cytokine were IL□6, 2□200 pg/mL; IL□10, 2□300 pg/mL; IL□17A, 2□1000 pg/ mL; TGF-β, 20 – 500 pg/mL.

### 2.4 Immunophenotyping

Whole blood samples (100 μL) were incubated with monoclonal surface antibodies (all anti-human) at 4 ºC for 20 min in accordance with the following combination: a) FITC-conjugated anti-CD4, Pe-conjugated anti-CD25, PerCP-Cy5.5-conjugated anti-CD39, APC-conjugated anti-CD73; b) FITC-conjugated anti-CD8, Pe-conjugated anti-CD39, APC-conjugated anti-CD73; c) PerCP-Cy5.5-conjugated anti-CD19, Pe-conjugated anti-CD39, APC-conjugated anti-CD73; d) FITC-conjugated anti-CD8, Pe-conjugated anti-CD28, PerCP-Cy5.5-conjugated anti-CD27, APC-conjugated anti-PD-1. Then, samples were incubated for 10 min with lysing buffer (BD Biosciences) and then centrifuged at 500 g for 5 min. The supernatant was discarded, and samples were resuspended in phosphate buffered saline (PBS 1 mL, pH 7.2) and then centrifuged at 500 g for 5 minutes. Finally, the samples were resuspended in 0.5 mL PBS and analyzed in flow cytometry. Cell phenotype was acquired using CELLQuest Pro Software (BD Bioscience) on a FACSCalibur flow cytometer (BD Bioscience). A minimal of 50.000 events/tubes were acquired, and granulocytes, lymphocytes and monocytes were identified and gated according to each forward scatter (FSC) and side scatter (SSC) profiles. In the CD8+ T cells gate, cells were further characterized by CD27/CD28 expression: CD27+CD28+ were defined as low-differentiated cells, and CD27-CD28-were defined as high-differentiated cells (18). PD-1 expression was evaluated in both CD8+CD27-CD28- and CD8+CD27+CD28+ T cells subsets and presented as mean fluorescence intensity (MFI). Supplementary Figure 1 shows the gate strategy for each analysis.

### 2.5 Mitochondrial membrane polarization and apoptosis analysis

The mitochondrial membrane potential (ΔΨm) was quantified according to a method previously described (19), using the fluorescent dye rhodamine 123 (Rh 123, Sigma-Aldrich). The detection of apoptosis was performed by using additional labeling with Annexin V-FITC following manufacturer’s guidelines (Invitrogen, ThermoFisher, USA). Analyses were performed by using CELLQuest Pro Software (BD Bioscience) on a FACSCalibur flow cytometer (BD Bioscience).

### 2.6 Cell culture and in vitro experiments

Peripheral blood mononuclear cells (PBMC) of a healthy non-infected donor were isolated from peripheral blood using histopaque gradient solution, Histopaque 1077 (Sigma□Aldrich, St Louis, MO, USA) as previously described (20). Then, the cells were washed and suspended in Roswell Park Memorial Institute□1640 medium (Sigma□Aldrich, USA) supplemented with 2 g/L sodium bicarbonate, 2% glutamine, 100 U/mL penicillin – 0.1 mg/mL streptomycin (Sigma□Aldrich, USA) and 10% of plasma acquired from mild COVID-19 (n=9), severe COVID-19 (n=9) or healthy controls (n=9) for 15 hours (37 ºC, 5% CO_2_). PBMCs were collected, washed with PBS 1x, and stained with anti-CD4 Pe, anti CD8 FITC, anti-CD39 Percp-Cy5.5 and anti-CD73 APC (all anti-human antibodies from BD Bioscience) for Flow Cytometry analysis in BD FacsCalibur (BD, USA).

To test the effect of *in vitro* adenosine treatment on COVID-19-related hyperinflammation, PBMC (1×10^6^ cells/well) of severe (n=3) COVID-19 patients were seeded in 12 well plates and treated with adenosine (100 μM) or PBS as control for 24 hours (37 ºC, 5% CO_2_). Then, cells were stained with 5 μL monoclonal antibodies (all anti-human) conjugated with specific fluorochromes: CD3 FITC (EbioScience, USA) or CD14 FITC (BIOGEMS, USA). Thereafter, cells were washed with 1.5 mL PBS. For intracellular analysis, cells stained with anti-CD3 or anti-CD14 antibodies were incubated with fixation and permeabilization buffers following manufacturer’s recommendation (Ebioscience, EUA). After the period, samples were incubated with Phospo-NF-κBp65 (Ser529) Pe (Ebioscience, EUA) monoclonal antibody during 30 min. Then, cells were washed and resuspended in Flow Cytometry Staining Buffer (Ebioscience, EUA) and analyzed by flow cytometry as described above.

### 2.7 Statistical analysis

Normality of data was checked by Kolmogorov□Smirnov, and the values were presented as mean ± standard deviation (SD). Categorical variables were presented as relative frequency and analyzed by Qui-Square test. To identify associations between adenine-based purine levels and symptoms we used Mann-Whitney U-test to compare SARS-CoV-2 infected patients with and without specific disease symptoms. A one□way ANOVA followed by Bonferroni’s post□hoc for multiple comparisons was used to verify between-groups differences. Pearson’s Coefficient Test was performed to check correlations between the variables. P value ≤ 0.05 were considered statistically significant. The SPSS 20.0 (IBM Inc, EUA) software was used in all analysis.

## 3. Results

### 3.1 Participant’s characteristics

The characteristics of severe (n=22), mild (n=24) COVID-19 patients, and healthy control (n=13) participants are presented in Table 1. All patients had COVID-19 confirmed by RT-PCR. Severe COVID-19 patients were older (p<0.001), presented more days from symptom onset (p<0.001) and required oxygen use during hospitalization (p<0.001) compared to mild COVID-19 patients. Furthermore, several symptoms were reported by severe COVID-19 patients, including cough (p=0.007), dysgeusia (p=0.04), anosmia (p=0.001), vomiting (p=0.007), diarrhea (p=0.004), skin rash (p=0.01), fatigue (p=0.01) and stuffy nose (p=0.005). Regarding associated medical condition, severe COVID-19 patients reported hypertension (p=0.002), diabetes mellitus (p=0.001), cardiovascular diseases (p=0.001), heart failure (p=0.001), chronic obstructive pulmonary disease (p=0.001), asthma (p=0.001) and dyslipidemia (p=0.001). (Table 1).

**Table 1.** Demographic and clinical characteristics of participants

|  | Healthy controls<br>(n=13) | Mild COVID-19<br>(n=22) | Severe COVID-19<br>(n=24) | p |
| --- | --- | --- | --- | --- |
| Age (y) | 52.3 ± 18.5 | 41.0 ± 10.6 | 60.0 ± 17.2 | **<0.001** |
| Sex (male, %) | 23.1 | 30.4 | 59.1 | 0.055 |
| **Racial or ethnic group (%)** |  |  |  |  |
| Caucasian | 75 | 69.6 | 77.3 | 0.34 |
| Non-caucasian | 25 | 30.4 | 22.7 |  |
| **Height (m)** |  | 1.68 ± 0.1 | 1.68 ± 0.0 | 0.98 |
| **Body mass (kg)** |  | 78.0 ± 18.8 | 82.1 ± 14.5 | 0.42 |
| **BMI (kg/m<sup>2</sup>)** |  | 27.3 ± 4.7 | 28.6 ± 4.0 | 0.33 |
| **BMI (%)** |  |  |  |  |
| Lean |  | 43.5 | 23.8 | 0.38 |
| Overweight |  | 26.1 | 38.1 |  |
| Obese |  | 30.4 | 38.1 |  |
| **Days from symptom onset to sample collection** | - | 3.4 ± 1.6 | 9.9 ± 3.8 | **0.001** |
| **Symptoms (%)** | - |  |  |  |
| Fever | - | 47.8 | 63.6 | 0.26 |
| Coryza |  | 47.8 | 54.5 | 0.65 |
| Cough |  | 65.2 | 95.5 | **0.007** |
| Anosmia |  | 21.7 | 45.5 | 0.10 |
| Dysgeusia |  | 26.1 | 45.1 | **0.04** |
| Dyspnea |  | 21.7 | 77.3 | **0.001** |
| Sore throat |  | 52.2 | 45.5 | 0.23 |
| Headache |  | 69.6 | 68.2 | 0.23 |
| Nausea |  | 17.4 | 40.9 | 0.06 |
| Vomiting |  | 0 | 22.7 | **0.007** |
| Diarrhea |  | 17.4 | 63.6 | **0.004** |
| Conjunctivitis |  | 30.4 | 31.8 | 0.26 |
| Skin rash |  | 0 | 9.1 | **0.01** |
| Fatigue |  | 52.2 | 86.4 | **0.01** |
| Myalgia |  | 60.9 | 72.7 | 0.17 |
| Chills |  | 34.8 | 68.2 | 0.053 |
| Appetite loss |  | 43.6 | 72.7 | 0.08 |
| Sputum production |  | 30.4 | 40.9 | 0.11 |
| Stuffy nose |  | 60.9 | 22.7 | **0.005** |
| **Underlying medical condition (%)** |  |  |  |  |
| Hypertension |  | 4.3 | 40.9 | **0.002** |
| Diabetes Mellitus |  | 0 | 22.7 | **0.001** |
| Cardiovascular diseases |  | 0 | 9.1 | **0.001** |
| Heart Failure | - | 0 | 4.5 | **0.001** |
| COPD |  | 4.3 | 9.1 | **0.001** |
| Asthma |  | 0 | 9.1 | **0.001** |
| HIV |  | 4.3 | 0 | **0.001** |
| Dyslipidemia |  | 0 | 9.1 | **0.001** |
| Oxygen use during hospitalization (%) | - | 0 | 52.4 | **0.001** |

Quantitative data presented as mean ± standard deviation. Groups comparison by one way ANOVA ( $p < 0.05$ ).
Categorical variables were presented as relative frequency and analyzed by Qui-Square test.
BMI, body mass index; COPD, chronic obstructive pulmonary disease; HIV human immunodeficiency virus.
Qualitative data presented as relative frequency. Between groups comparison through Qui-square test ( $p < 0.05$ ).

### 3.2 COVID-19 impacts on systemic purinergic molecules and cytokine levels

The plasma concentrations of adenine-based purines ATP, ADP, AMP, adenosine and cytokines are shown in Figure 1. ATP levels were decreased in mild (p=0.03) and severe (p=0.03) COVID-19 patients compared to healthy controls. Systemic adenosine levels were higher in healthy controls compared to mild (p=0.03) and severe (p=0.02) COVID-19 patients. Severe COVID-19 presented higher systemic IL-6 and IL-10 levels compared to healthy controls and mild COVID-19 patients (p<0.01 for all comparisons). In addition, lower IL-17A levels were identified in the peripheral blood of healthy controls compared to mild (p=0.03) and severe COVID-19 (p=0.02) patients. Lower frequencies of CD4+ T cells (p<0.001 vs. healthy controls; p=0.03 vs. mild COVID-19) and CD8+ T cells (p<0.001 vs. healthy controls) were found in severe COVID-19 patients. Mild COVID-19 patients also presented lower proportions of CD8+ T cells compared to healthy patients (p=0.02). Reduced peripheral frequency of CD4+CD25-T cells were also found in mild COVID-19 (p=0.01) and severe COVID-19 (p=0.001) groups compared to healthy controls. No differences were identified in ADP, AMP, TGF-β levels and in CD19+ B cell frequency among the groups (p>0.05).

**Figure 1.**
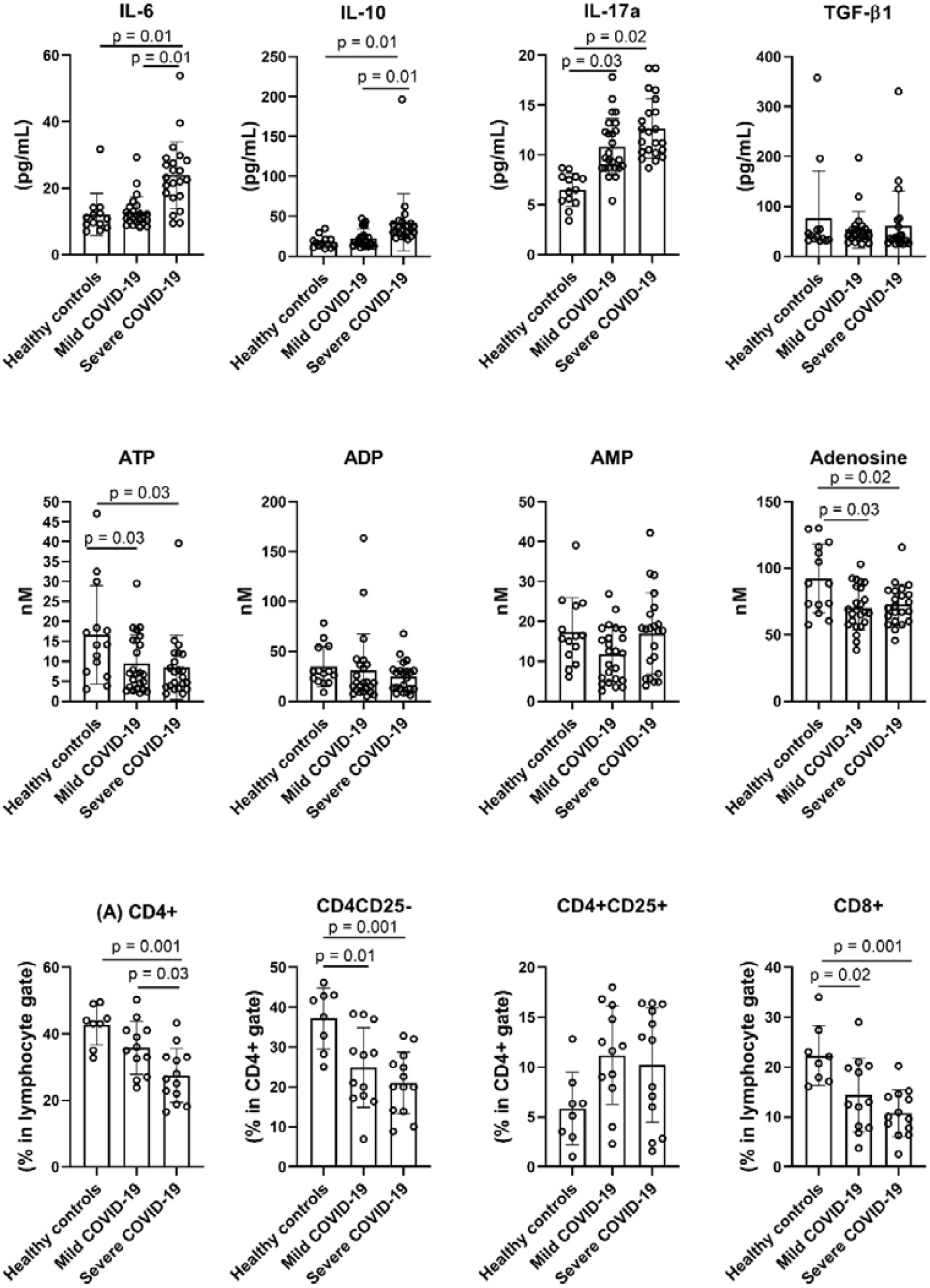
The systemic levels of systemic cytokines, adenine-based purines, and the peripheral frequency of CD4+, CD4+CD25-, CD4+CD25+, and CD8+ T cells of healthy controls, mild and severe COVID-19 patients. Data are presented as mean ± standard deviation. Group comparisons were performed by one-way ANOVA with Bonferroni’s post-hoc (p≤0.05).

Then, we compared the systemic levels of adenine-based purines in patients with or without clinical symptoms of COVID-19. COVID-19 patients with nausea symptom (n=13) presented lower adenosine levels (p=0.04) and a tendency to decreased AMP levels (p=0.07). Furthermore, higher ADP levels were found in patients with stuffy nose (p=0.02, n= 19). Skin rash was reported by two patients, and presented lower ATP (p=0.01), ADP (p=0.01) and AMP (p=0.01) levels (Figure 2). Interestingly LPS levels, a microbial translocation marker who is elevated in COVID-19 patients, were higher in patients with nausea symptom (p=0.02) and inversely correlated with the systemic adenosine levels (r= −0.54; p= 0.03).

**Figure 2.**
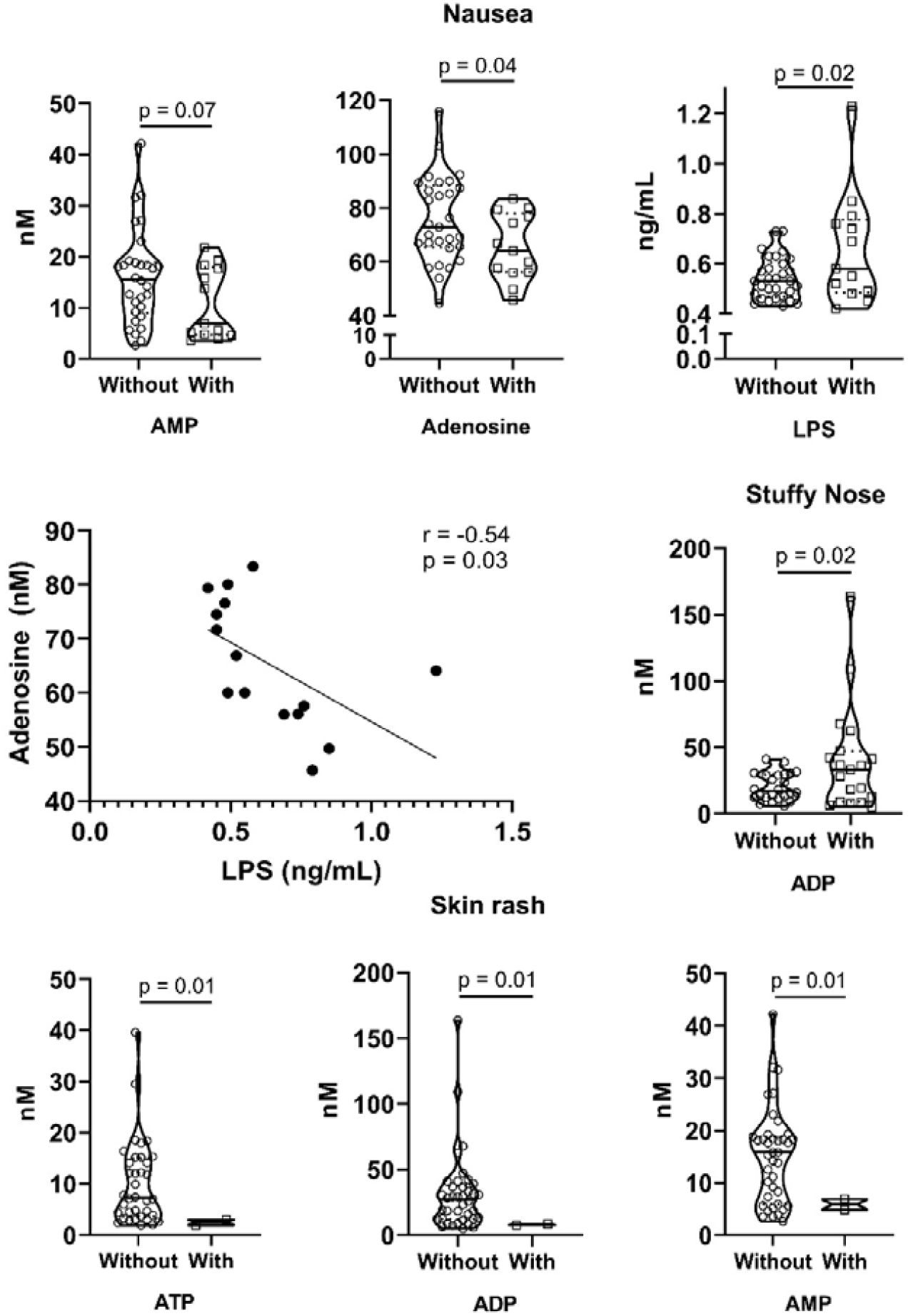
The association of ATP, ADP, AMP and adenosine levels, and LPS concentrations with clinical symptoms of acute SARS-CoV-2 infection.

The levels of adenine-based purine molecules and endotoxin were compared between patients with and without clinical symptoms through Mann-Whitney U-test and the significant results were presented (p<0.05). The correlation between adenosine levels and LPS concentrations in patients reporting nausea symptom were performed by Pearson’s Coefficient Correlation test (p<0.05).

### 3.2 Altered CD39 and CD73 expression in T lymphocytes of COVID-19 patients

The T cell phenotype was evaluated in healthy controls (n=8), mild COVID-19 (n=12) and severe COVID-19 patients (n=13). The peripheral frequencies of CD4+, CD4+CD25- and CD4+CD25+ T cells expressing CD39+ and CD73+ ectonucleotidases were presented in Figure 3. In total CD4+ T cells, severe COVID-19 group had higher frequencies of CD4+CD39+CD73-(p=0.01 vs. healthy controls; p=0.03 vs. mild COVID-19; Fig.3A), but lower CD4+CD39-CD73+ (p=0.04 vs. healthy controls; Fig.3B). The peripheral frequency of CD4+CD25-CD39+CD73-was higher in severe COVID-19 (p=0.002 vs. healthy controls; p=0.004 vs. mild COVID-19; Fig.3D), without differences in the CD4+CD25-CD39-CD73+ or CD4+CD25-CD39+CD73+ T cells proportions (p>0.05; Fig.3E-F). Healthy controls had higher proportions of CD4+CD25+CD39+CD73-(p=0.002 vs. mild COVID-19; p=0.001 vs. severe COVID-19; Fig.3G) and CD4+CD25+CD39+CD73+ (p=0.001 vs. mild COVID-19; p=0.001 vs. severe COVID-19; Fig.3I).

**Figure 3.**
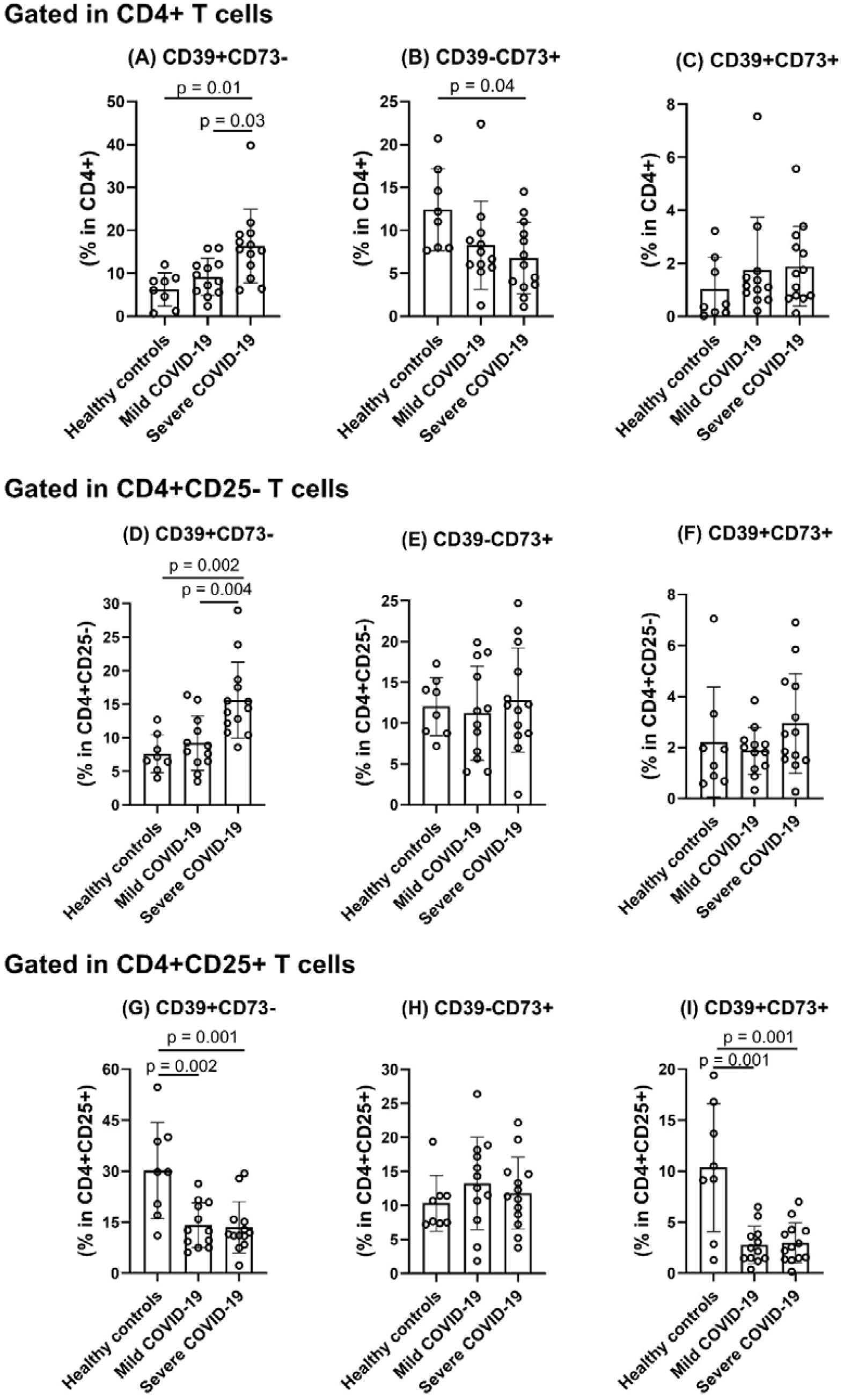
The frequency of CD4+ T cells expressing CD39 and CD73 ectonucleotidases in the peripheral blood of healthy controls, mild and severe COVID-19 patients. Data are presented as mean ± standard deviation. Group comparisons were performed by one-way ANOVA with Bonferroni’s post-hoc (p≤0.05).

Figure 4 shows the CD8+ T cell phenotype in COVID-19 patients and healthy controls. CD8+CD39-CD73+ T cells were lower in severe COVID-19 (p=0.001 vs. healthy controls; p=0.04 vs. mild COVID-19; Fig.4B). Higher frequency of high-differentiated (CD27-CD28-) CD8+ T cells (Fig.4D) was identified in the peripheral blood of mild COVID-19 patients (p=0.001 vs. healthy control) and severe COVID-19 subjects (p=0.001 vs. healthy control), and lower proportions of low-differentiated (CD27+CD28+) CD8+ T cells in the severe COVID-19 group (p=0.05 vs. healthy controls, Fig.4E). The expression of PD-1 was higher in low-differentiated CD8+ T cells of severe COVID-19 patients (p=0.008 vs. healthy controls, Fig.4G).

**Figure 4.**
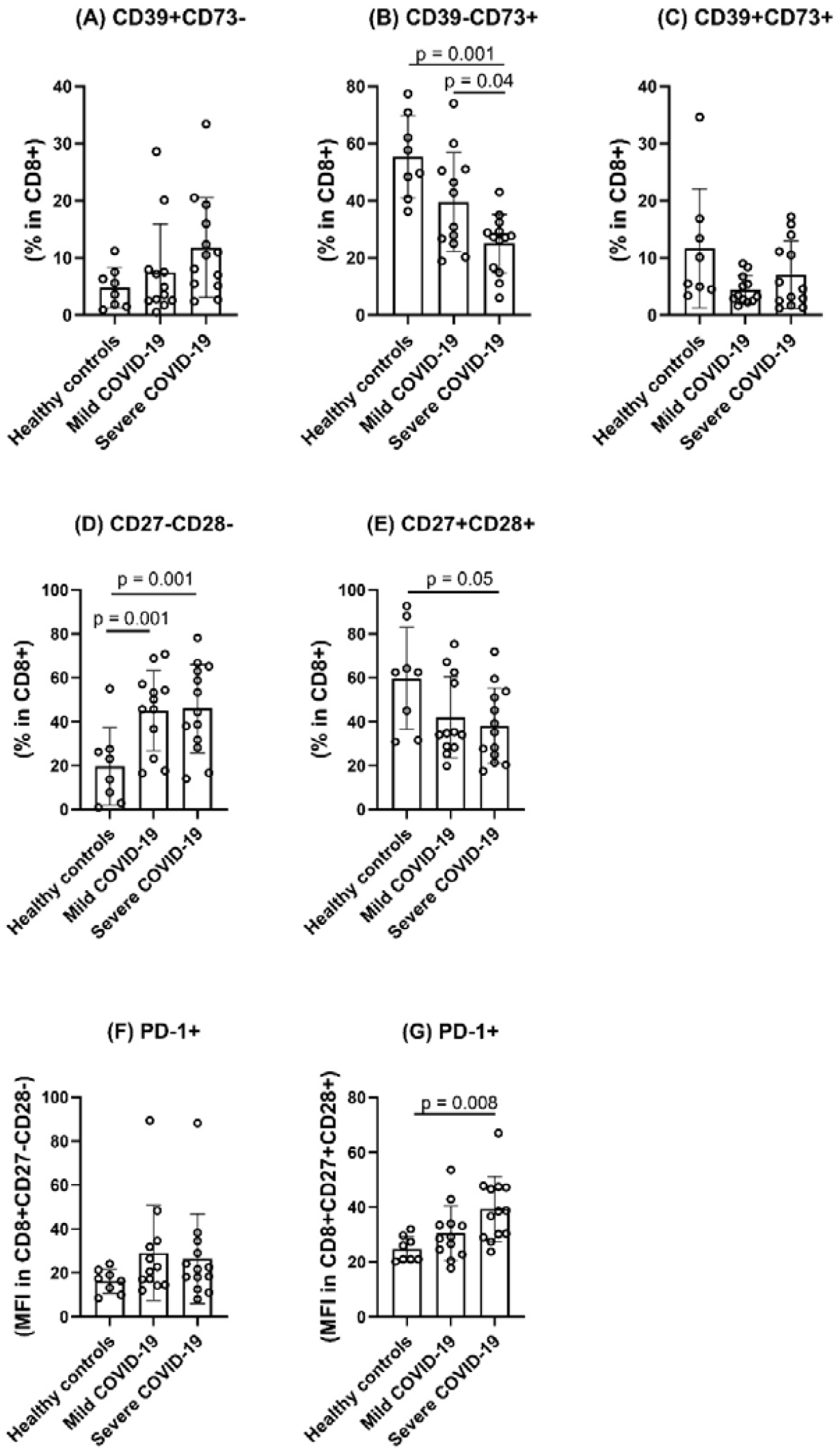
The frequency of CD8+ T cells expressing CD39, CD73, CD27, CD28 and PD-1 in the peripheral blood of healthy controls, mild and severe COVID-19 patients. Data are presented as mean ± standard deviation. Group-comparisons were performed by one-way ANOVA with Bonferroni’s post-hoc (p≤0.05).

### 3.3 Higher apoptosis and mitochondrial dysfunction in lymphocytes of COVID-19 patients

Next, we evaluated the apoptosis and mitochondrial membrane polarization in healthy controls (n=6), mild COVID-19 (n=6) and severe COVID-19 (n=6) individuals (Figure 5). COVID-19 patients presented decreased mitochondrial membrane polarization (severe COVID-19 vs. healthy control, p<0.001; mild vs. healthy control, p=0.03; Fig.5A), indicating mitochondrial membrane depolarization. COVID-19 patients presented higher CD4+Annexin V+ (severe COVID-19 vs. healthy controls, p=0.001; mild COVID-19 vs. healthy controls, p=0.01) and CD8+Annexin V+ (severe COVID-19 vs. healthy controls, p=0.001; mild COVID-19 vs. healthy controls, p=0.01). Severe COVID-19 patients also presented higher CD8+Annexin V+ T cells compared to mild COVID-19 subjects (p=0.03). (Fig.5B).

**Figure 5.**
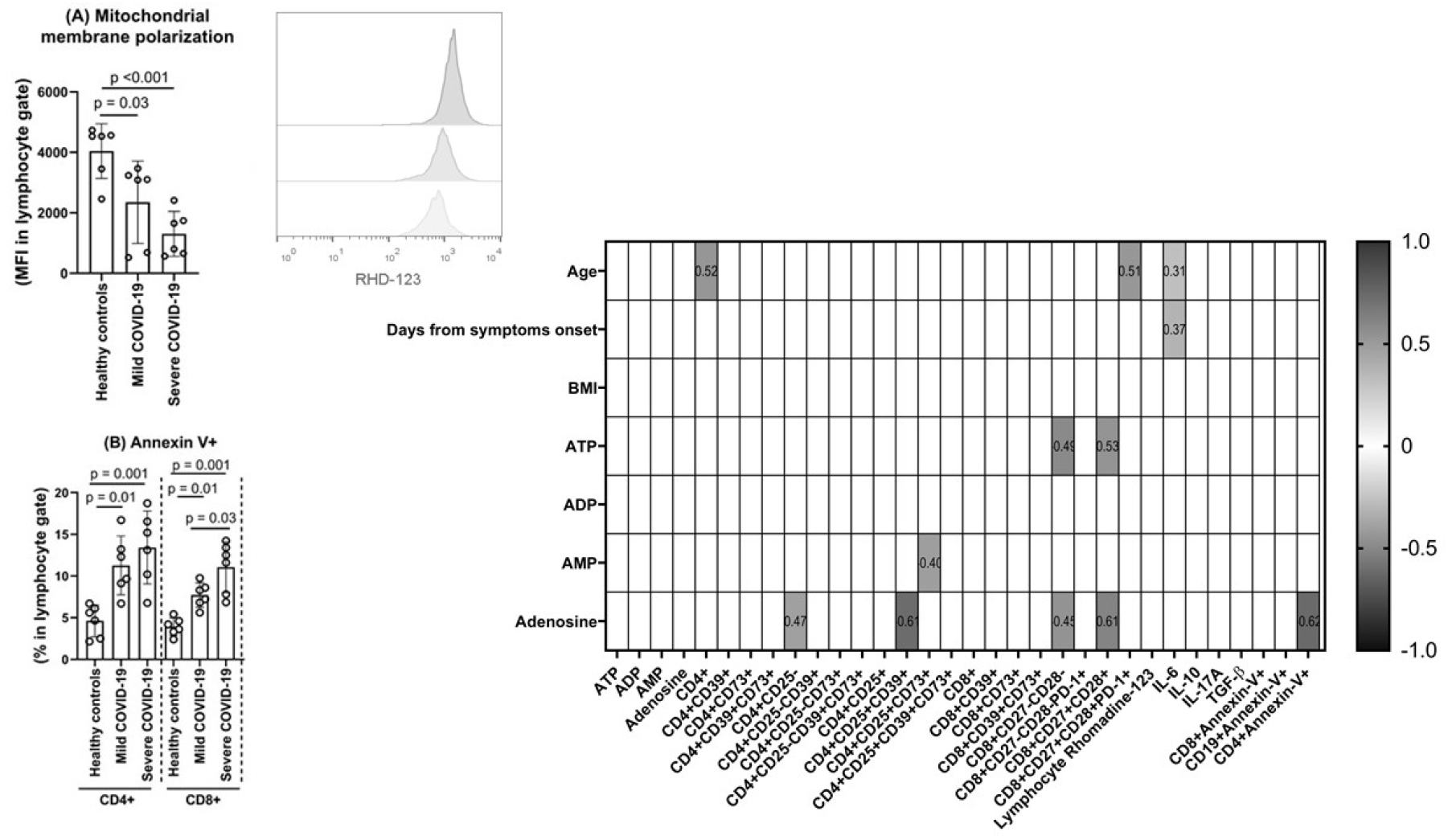
Mitochondrial membrane polarization (Fig.5A) and apoptosis (Fig.5B) of lymphocytes in COVID-19 patients and healthy controls. Data are presented as mean ± standard deviation. Group-comparisons were performed by one-way ANOVA with Bonferroni’s post-hoc (p≤0.05).

Figure 5C shows the heat map of Pearson’s Coefficient Correlation Test. ATP levels inversely correlated with CD8+CD27-CD28- (r=-0.49; p=0.01) and positively correlated with CD8+CD27+CD28+ T cell proportions (r=0.53; p=0.03). AMP levels inversely correlated with CD4+CD25+CD73+ (r=-0.40; p=0.04). Adenosine levels positively correlated with the peripheral frequency of CD4+CD25- (r=0.47; p=0.01) and CD8+CD27+CD28+ (r=0.61; p=0.001), but negatively correlated with CD4+CD25-CD39+ (r=-0.61; p=0.001), CD4+CDCD27+CD28+ (r=-0.45; p=0.02) and CD4+Annexin-V+ (r=-0.62; p=0.03) proportions. The age of the COVID-19 patients correlated with the frequency of CD4+ T cells (r=0.52; p=0.03), the expression of PD-1+ on CD8+CD27+CD28+ T cells (r=0.51; p=0.02) and the IL-6 levels (r=0.31; p=0.04). Days from symptoms onset correlated only with the plasma IL-6 levels (r=0.37; p=0.02).

The Correlation heat map including adenine-based purines and immune variables at admission to the Hospital (Fig. 5C). Pearson’s correlation coefficients are plotted. Cells were colored according to the strength and trend of correlations (shades of red=positive, shades of blue=negative correlations). Statistical analysis performed through Pearson’s coefficient correlation.

### 3.4 In vitro adenosine treatment reduces NF-_κ_B expression in PBMC

The incubation of PBMC from a healthy non-infected control donor with the plasma of severe COVID-19 patients decreased the expression of CD73 on the cell surface of CD4+ (Fig.6A; p=0.01 vs. healthy controls; p=0.02 vs. mild COVID-19) and CD8+ (Fig.6B; p=0.01 vs. healthy controls; p=0.02 vs. mild COVID-19) T cells, without changes in CD39 expression. Interestingly, PBMC from severe COVID-19 patients *in vitro* treated with adenosine (100 μM) reduced the NF-κB activation in both CD3+ T cells (Fig.6C, p=0.04) and CD14+ monocytes (Fig.6D, p=0.04).

**Figure 6.**
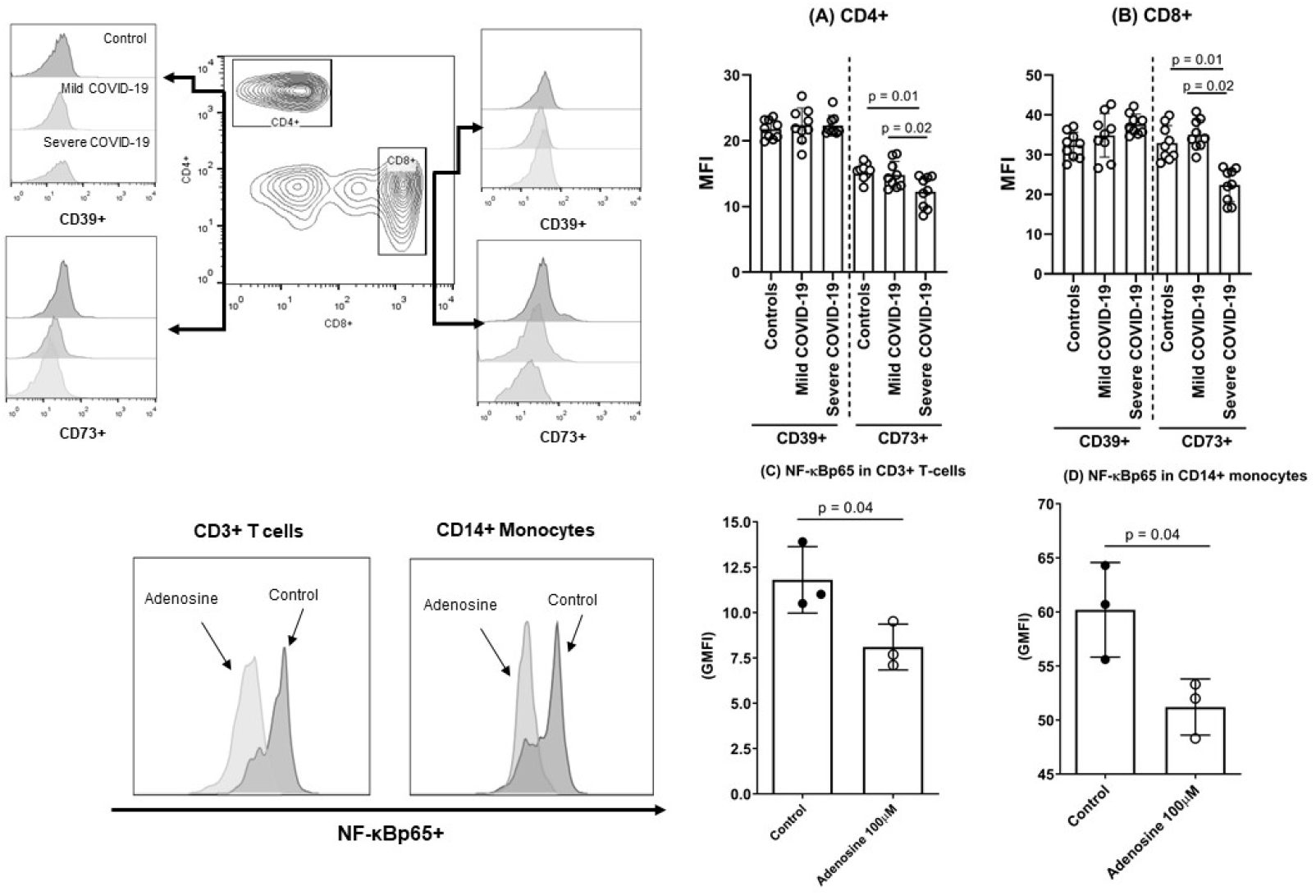
CD39 and CD73 expression in CD4+ (Fig.6A) and CD8+ (Fig.6B) T cells of a healthy noninfected donor after the incubation with plasma obtained from controls, mild COVID-19 or severe COVID-19 patients. In addition, PBMC of severe COVID-19 (n=3) were treated *in vitro* with adenosine and the activation of NF-κBp65 were evaluated in CD3+ T cells (Fig.6C) and CD14+ monocytes (Fig.6D).

Data are presented as mean ± standard deviation. Group-comparisons were performed by one-way ANOVA with Bonferroni’s post-hoc (p≤0.05).

## 4. DISCUSSION

This study showed for the first time the relationship between COVID-19 severity and lower systemic adenine-based purines and the frequency of circulating lymphocytes expressing CD39 and CD73 ectonucleotidases. We found decreased plasma levels of ATP and adenosine in COVID-19 patients, concomitant with markedly altered frequencies of CD4+ and CD8+ T cells expressing CD39 and CD73 enzymes on the cell surface. In addition, plasma samples collected from severe COVID-19 patients were able to decrease the expression of CD73 on CD4+ and CD8+ T cells of healthy donor and the *in vitro* incubation of PBMC from severe COVID-19 patient with adenosine reduced the NF-κB activation in T cells and monocytes. Furthermore, COVID-19 severity was associated with an accumulation of high differentiated CD27-CD28-CD8+ T cells proportions and decreased low-differentiated CD27+CD28+ CD8+ T cell frequency in the peripheral blood concomitant with higher expression of the exhaustion marker PD-1 in this population. COVID-19 patients also presented higher rates of lymphocytes mitochondrial membrane depolarization and increased apoptosis rates in CD4+ and CD8+ T cells.

Lower ATP plasma levels is a surprising new result in SARS-CoV-2 infection. Extracellular ATP was found at high concentrations in bronchoalveolar lavage fluid (BALF) from patients with acute respiratory distress syndrome and in the blood of septic patients (21). On the other hand, extracellular ATP facilitates IFN type I secretion through p38/JNK/ATF-2 signaling pathway in vesicular stomatitis virus (VSV)-infected mice (22). Thus, limiting the extracellular levels of ATP in SARS-CoV-2 infection may contribute to the delay in IFN type I, turning off the initial alarm of the purinergic system. SARS-CoV-2 infection impairs the rapid IFN-I pathway activation in mild and severe patients, leading to lower antiviral response and worse outcomes in critically ill COVID-19 patients (23). Also, the correlation analysis revealed a positive correlation between extracellular ATP levels and the peripheral frequency of low differentiated CD8+ T cells. This result may suggest that decreased extracellular ATP during COVID-19 disease may impact the regulation of total CD8+ T cell differentiation.

Here, for the first time we showed that COVID-19 patients present lower adenosine levels in the blood, which may contribute to the uncontrolled inflammation, and severity of disease. Adenosine binds the P1 receptors, mainly the AR2A subtype, of immune cells (i.e. lymphocytes, monocytes and macrophages) to induce several events to suppress the inflammatory response, such as the downregulation of the master inflammatory transcription factor nuclear factor kappaB (NF-kB) (8,24,25). Furthermore, our data revealed higher levels of IL-6, IL-10 and IL-17A plasma concentrations in severe COVID-19 group, indicating a Th1/Th17 inflammatory polarization. Acute hypercytokinemia is a common feature of severe COVID-19 that may contribute to the multiorgan impairment and coagulopathy disorders (17,26,27). Reinforcing the regulatory effect of adenosine in the inflammation and cytokine storm, the use of inhaled adenosine in COVID-19 patients has successfully decreased the length of hospitalization and improved the prognosis of patients (28,29). We found that in vitro adenosine treatment lowers the NF-κB activation in both T cells and monocytes, suggesting a role for the therapeutic adenosine treatment for the reduction of hyper inflammation in COVID-19. Collectively, our data indicate alterations in adenine-based purinergic molecules, mainly ATP and adenosine levels, during the initial phase of SARS-CoV-2 infection.

Here, we also described a negative correlation between plasma adenosine levels and the CD4+ T cell apoptosis rate. Previous studies have suggested that adenosine and its analog are related to anti-apoptotic events through A2A receptor activation in CD4+ T cells (30). On the other hand, mitochondrial dysfunction is involved in the induction of apoptosis, thus increasing the depolarization of transmembrane potential (31). Here, lymphocytes of COVID-19 patients presented a state of mitochondrial membrane depolarization and higher rates of apoptosis. Interesting, mitochondria have a central role in T cell activation by producing ATP, and higher mitochondrial dysfunction may leads to failure in the purinergic regulation and cell homeostasis (32).

The increased expression of CD39 and CD73 in lymphocytes rapidly converts extracellular ATP to adenosine to induce anti-inflammatory effects (4). Here, we described increased proportions of CD4+CD39+CD73-T cells and lower frequencies of CD4+CD39-CD73+ and CD8+CD39-CD73+ in the peripheral blood of severe COVID-19 patients. CD39 cleaves ATP to ADP and AMP, which is hydrolyzed to adenosine by CD73. In this line, the higher expression of CD39 together with lower CD73 on T cells may explain the diminished extracellular levels of both ATP and adenosine concomitant to an exacerbated inflammatory state in COVID-19 patients.

CD39 was firstly described as an activation marker found in B cells, CD4+ and CD8+ T cells, CD14+ monocytes and NK cells (33). In COVID-19 patients, higher CD39+ T cells frequency was previously associated with the expression of PD-1, confirming the hypotheses that SARS-CoV-2 infection induces an hyperactivated exhausted lymphocyte profile (34), contributing to the T cell dysfunction in COVID-19. Furthermore, CD4+CD39+ T cells identifies an effector lymphocyte that are prone to apoptosis in older adults (35), corroborating with our data regarding higher apoptosis rate in CD4+ T cells of COVID-19 patients. On the other hand, lower frequencies of CD4+CD39-CD73+ and CD8+CD39-CD73+ T cells were identified in severe COVID-19 patients. These results are in line with some recent data that demonstrated an association between skewing of TCR repertoire with early CD4+ and CD8+ activation and altered expression of several immune checkpoints, such as Tim-3, PD-1, and CD73 (36). The downregulation of CD73 impairs not only the extracellular adenosine generation and anti-inflammatory response, but also the modulation of innate immune activation during the viral immune response (11). In severe COVID-19 patients, CD8+ T cells lacking CD73 expression possess a significantly cytotoxic effector functionality compared to CD8+CD73+ T cells, and correlates with plasma ferritin level, a surrounding clinical marker of uncontrolled systemic inflammation (13). Here, the diminished proportions of CD8+CD73+ T cells were accompanied by low frequencies of CD8+CD27+CD28+ low differentiated T cells and higher proportions of CD8+CD27-CD28-T cells in the peripheral blood of COVID-19 patients. Highly differentiated CD27-CD28-cytotoxic T cells presents low telomerase activity and decreased Akt phosphorylation which limits the ability of these cells to be maintained in continuous proliferation after antigen recognition (37). Although we were unable to determine the memory phenotype of T cells due to methodological reasons, our results indicate that COVID-19 patients display a complex alteration in differentiation status and memory acquisition in adaptive immunity related to disturbances in purinergic signaling. De Biasi and colleagues (15) showed that COVID-19 patients presented lower proportions of naive CD8+ T cells (CCR7+CD45RA+CD28+CD27+) and increased frequencies of terminally effector CD8+ T cells (CCR7-CD45RA+CD28-CD27-) compared to healthy patients. Moreover, we found increased PD-1 expression in low differentiated CD8+CD27+CD28+ T cells of severe COVID-19 group, indicating an exhaustion profile.

The combining analysis of CD25 and CD39 expression in CD4+ T cells reveals distinct phenotypes: CD4+CD25+CD39+ (activated/memory Treg, mTreg); and CD4+CD25−CD39+ (memory T effector cell; mTeff) (38,39). These populations dynamically change during the acute inflammatory response in response to extracellular purine molecules accumulation (40). We found an accumulation of CD4+CD25-CD39+ mTeff cells in severe COVID-19 patients. CD39+ mTeff cells is non-suppressive and has a memory phenotype that express higher levels of mRNA Th lineage specific cytokine profile, mainly Th1 and Th17 subsets (41). In this sense, the accumulation of CD4+CD25-CD39+ T cells in the peripheral blood of severe COVID-19 patients may contribute to the skewing toward Th17 profile. In humans CD4+CD25-CD39+ T cells are CD45RO+, Foxp3-, secrete higher amounts of IFN-γ and IL-17 and are increased following autoantigen recognition (42). Thus, the upregulation of CD39 expression in CD4+CD25-T cells is a consistent consequence of activating T helper cells after tissue injury and inflammatory exacerbation.

CD4+CD25+CD39+ mTreg cells were decreased in mild and severe COVID-19 patients. Furthermore, the proportions of CD4+CD25+ T cells co-expressing CD39 and CD73 were also reduced in COVID-19 patients. These data indicate a failure in the immunosuppressive axis of adenosine generation by Treg cells during SARS-CoV-2 infection. Recently, Meckiff and coworkers (43) described an imbalance of regulatory T cells (Tregs) and cytotoxic reactive CD4+ T cells in severe COVID-19. Since Treg cells are known to play an important role in limiting the host antiviral response and the consequent tissue immunopathology, the present data showing decreased CD39/CD73 axis in Treg that might decrease Treg response to SARS-CoV-2 and have a relevant impact on fueling systemic inflammation in COVID-19 patients. (44).

## 5. CONCLUSION

In summary, we describe for the first time an imbalance in the levels of extracellular adenine-based purine molecules and alterations in CD39/CD73 ectonucleotidases axis in CD4+ and CD8+ T cells of COVID-19 patients with different degrees of disease severity. The reduced adenosine extracellular levels and alterations in T cells phenotype may impact on COVID-19 severity. Collectively, these data add new knowledge regarding the immunopathology of COVID-19 through purinergic regulation.

## Data Availability

Data will be available when requested.

## Acknowledgments

We are grateful to the Brazilian agencies Coordenação de Aperfeiçoamento de Pessoal de Nível Superior (CAPES) – Finance Code 001. This study was supported in part by the Hospital Moinhos de Vento through the Program for Supporting the Institutional Development of the Public Health System (PROADISUS), supported by the Ministry of Health of Brazil. GPD is supported by postdoctoral *fellowship from* CAPES. PRTR, CB and CRB are grateful to *CNPq* for the *PQ productivity scholarship*. Finally, we thank all subjects who agreed to participate in the study and donate blood.

